# Barriers and facilitators to Medical Emergency Team activation among healthcare professionals: protocol for a multicentre cross-sectional survey in four Swiss hospitals

**DOI:** 10.64898/2026.09.01.26359689

**Authors:** Nicole Goetz, Aline Wolfensberger, Roger Ludwig, Joerg C. Schefold, François Fontana, Andrea R Kopp Lugli, David Berger, Daniel Tuchscherer, Raoul Suter, Luca Cioccari

## Abstract

**Background:** Medical Emergency Teams (METs) are intended to provide early expert assessment and treatment for deteriorating hospitalised patients. However, MET activation may occur only after prolonged physiological deterioration. Potential explanations include uncertainty about activation criteria, professional hierarchy, departmental culture, perceived pressure to manage deterioration without help, fear of criticism, and concerns that activation may reflect poorly on professional competence. This study aims to identify modifiable barriers and facilitators to timely MET activation among healthcare professionals caring for hospitalised adults.

**Methods:** This multicentre cross-sectional web survey will be conducted sequentially at four Swiss hospitals: two cantonal hospitals and two tertiary university hospitals. Eligible participants are physicians, nurses, nurses in training, physiotherapists, occupational therapists, and speech and language therapists involved in the care of hospitalised adult patients in areas where MET activation may occur. Staff working primarily in paediatrics, emergency medicine, anaesthesia, intensive care, operating theatres, recovery areas, or outpatient care, and MET members themselves, are excluded. The German-language questionnaire was developed from relevant items identified in published MET and rapid response system surveys and was informed by the revised 14-domain Theoretical Domains Framework (TDF). The TDF was used to assess the theoretical breadth of the candidate item pool and to ensure coverage of all domains judged relevant to timely MET activation. Two physician investigators independently mapped candidate items to the single TDF domain judged most appropriate; disagreements were resolved by consensus. All 14 TDF(v2) domains were judged potentially relevant to the target behaviour, and additional items were developed when relevant domains were not adequately represented. The questionnaire was pilot tested by physicians and nurses. The four prespecified primary outcomes comprise one proximal self-reported behavioural indicator - non-activation despite fulfilment of MET criteria - and three potential determinants of timely activation: perceived pressure to avoid activation, concern that activation may reflect poorly on professional competence, and absence of a departmental culture supporting immediate activation. These outcomes were specified in the protocol and statistical analysis plan before inspection of the pooled multicentre results. The primary professional comparison is physicians versus nurses. Ordinal logistic regression will adjust for hospital, years of professional experience, clinical area, and previous personal MET activation. Secondary outcomes will be analysed descriptively and exploratorily. No comparison by MET maturity will be undertaken.

**Ethics and dissemination:** The project was classified as research outside the scope of the Swiss Human Research Act. Site-specific ethics determinations and institutional endorsements will be reported. Participation is voluntary. Responses are confidential rather than fully anonymous because optional contact details may be entered in the same survey record. Results will be disseminated through a peer-reviewed publication, presentations, and aggregated feedback to participating hospitals.

**Registration:** Because data collection has already been completed at one hospital, the registration will be described as a prospective registration of the pooled analysis rather than a fully prospective preregistration.

## Introduction

Rapid response systems are designed to identify and treat deteriorating hospitalised patients before preventable cardiac arrest, unplanned intensive care admission, or other serious adverse events occur. The afferent component of the system depends on ward-based staff recognising deterioration and activating the Medical Emergency Team (MET), while the efferent component provides urgent expert assessment and treatment.

A central limitation of rapid response systems is afferent-limb failure: activation may be absent or delayed despite the presence of predefined physiological criteria or clinical concern. In the setting motivating this study, more than 50% of patients for whom the MET was activated had documented signs fulfilling MET criteria during the preceding 24 hours. This suggests that many calls may occur later than intended.

Previous survey research has identified barriers such as contacting the responsible ward physician before the MET, reluctance to activate when criteria are fulfilled but the patient does not appear critically ill, fear of criticism, concerns about professional competence, workload, and local cultural expectations. Conversely, previous MET experience, confidence, role clarity, supportive team culture, and perceived usefulness of the MET may facilitate activation.(1-3)

Implementation determinant frameworks can help organise these influences and identify potentially modifiable determinants of MET activation. The Theoretical Domains Framework (TDF) integrates constructs from multiple behavioural theories into 14 domains, including knowledge, skills, social/professional role and identity, beliefs about capabilities and consequences, intentions, environmental context and resources, social influences, emotion, and behavioural regulation.(4) The TDF can be used to examine barriers and facilitators to a clearly specified target behaviour and to inform subsequent intervention design.

## Objectives

The primary objective is to identify healthcare professionals’ reported barriers and facilitators to timely MET activation and to compare between physicians and nurses four prespecified primary outcomes: (1) self-reported non-activation despite fulfilment of MET criteria, a proximal behavioural indicator; (2) perceived pressure to avoid MET activation; (3) concern that MET activation may reflect poorly on professional competence; and (4) absence of a departmental culture supporting immediate MET activation. These outcomes were specified in the protocol and statistical analysis plan before inspection of the pooled multicentre results.

Secondary objectives are to:

- describe knowledge of the MET system, its purpose, activation process, and activation criteria;
- describe previous exposure to and personal activation of the MET;
- assess perceived usefulness, effectiveness, timeliness, and frequency of MET activation;
- assess confidence, communication, psychological safety, role clarity, and previous experiences with the MET;
- describe perceptions of local activation criteria and preferred activation approaches;
- identify improvement suggestions from free-text responses.

## Methods

### Study design and setting

This is a multicentre, cross-sectional, questionnaire-based study conducted sequentially at four Swiss hospitals: one tertiary cantonal hospital, one secondary cantonal hospital, and two tertiary university hospitals.

All hospitals operate an adult MET or equivalent rapid response system with broadly comparable purpose, availability, staffing, and activation pathways, although exact activation criteria differ slightly. The MET had been operational for approximately 1-2 years at the two cantonal hospitals and for more than 10 years at the two university hospitals. MET maturity is treated as contextual information only; no formal comparison according to implementation duration is planned.

The target behaviour is timely activation of the MET by healthcare professionals caring for a hospitalised adult patient who fulfils institutional activation criteria or whose condition causes serious clinical concern.

### Participants and eligibility

Eligible participants are physicians of all grades, registered nurses, nurses in training, physiotherapists, occupational therapists, and speech and language therapists involved in the care of hospitalised adult patients in clinical areas in which MET activation may occur.

Staff primarily caring for children, outpatients, emergency department patients, patients in operating theatres or recovery areas, or patients already admitted to intensive care are excluded. Anaesthesia and intensive care staff, MET members, and personnel not involved in care pathways where MET activation can occur are excluded. Radiology staff are eligible when they may encounter deterioration in hospitalised adult patients. Temporary, agency, locum, and rotating staff are eligible if they otherwise meet the criteria, although incomplete email coverage of these groups is anticipated. No minimum duration of employment is required.

Eligibility is communicated in the invitation and confirmed by an initial survey screening question. Respondents who do not confirm that they care for hospitalised adult patients in relevant clinical areas are exited from the survey and excluded from analysis.

### Questionnaire development

#### Literature review, TDF mapping, and item generation

A pragmatic literature review identified published questionnaires assessing attitudes, barriers, facilitators, and experiences related to METs and rapid response systems. Items judged relevant to the prespecified survey objectives were extracted from the identified questionnaires and compiled in a structured item bank; this did not imply automatic inclusion of every item from every source questionnaire. Published wording was retained whenever possible and adapted when required for Swiss clinical practice, local escalation pathways, or study-specific objectives.

Before developing additional items, the investigators considered each of the 14 TDF(v2) domains for its potential relevance to the specified target behaviour of timely MET activation. All 14 domains were judged potentially relevant. Candidate literature-derived items were then mapped to the TDF to assess the theoretical breadth of the existing item pool and identify relevant domains that were not adequately represented. New questions were developed to address these theoretical gaps and additional context-specific aspects of MET activation for which no suitable published item was available.

Questionnaire development was led by a senior intensivist and an intensivist in training in collaboration with an implementation scientist and an intensive care nurse. Published MET survey research informed items addressing, for example, professional hierarchy, fear of criticism, perceived competence, workload, criteria interpretation, perceived timeliness, learning, and communication.(1-3)

#### Use of the Theoretical Domains Framework

The revised 14-domain TDF, TDF(v2), was used as a theory-informed framework for questionnaire development and content coverage.(4) Two physician investigators independently assigned each candidate questionnaire item to the single TDF domain judged most appropriate. Multiple-domain allocation was not permitted. Disagreements were resolved through discussion and consensus; a third adjudicator was not required because consensus was reached for all items.

The purpose of the mapping was to assess theoretical coverage rather than to create psychometric domain scales. After the relevance of all 14 domains had been considered, the mapping exercise was used to determine whether the candidate item pool adequately represented each relevant domain. Additional items were developed for domains judged relevant but insufficiently represented and for system-specific topics. The final questionnaire therefore covers all 14 TDF(v2) domains. The questionnaire is considered TDF-informed, not a validated psychometric TDF instrument. Individual items, rather than TDF domain scores, are the units of analysis.

#### Linguistic and contextual adaptation

Candidate items originally published in English were linguistically and contextually adapted into German using DeepL and other artificial intelligence-assisted language tools. Each translation was independently reviewed and reconciled by the first and senior authors, both native German speakers and proficient in academic English.

All hospitals use the German questionnaire. The common core is identical across sites. Institution-specific terminology for the MET is retained to minimise confusion.

#### Pilot and usability testing

The final German questionnaire was pilot tested by physicians and nurses who were not eligible for the main survey. Participants completed the electronic questionnaire and provided written feedback. Minor linguistic revisions were made, and the revised questionnaire and branching logic were retested. Pilot responses are excluded from the study dataset.

### Questionnaire content

The questionnaire comprises 17 main numbered questions, several of which contain multiple statements or subitems. It includes eligibility screening, professional characteristics, previous MET exposure and activation, knowledge, activation attitudes, perceived departmental culture, benefits and concerns, psychological safety and communication, perceived timeliness, assessment of local criteria, preferred activation approaches, and free-text suggestions.

Most attitudinal items use a five-point Likert-type scale from strongly disagree to strongly agree. Other items use categorical, numerical, multiple-choice, or free-text formats. Adaptive branching displays follow-up items only when applicable.

### Recruitment and survey administration

Hospital executive leadership, including chief medical and chief nursing officers, and medical directors of participating departments endorsed the study before distribution.

Medical and nursing departmental leaders are asked by email to forward the survey invitation to all eligible staff and copy the research team on the forwarding message. Departments that do not confirm forwarding are contacted directly by the research team through available institutional email channels. Two reminders are distributed through departmental leadership or centrally by the research team.

The invitation explains the purpose, target population, approximate 10-minute completion time, voluntary nature, confidentiality arrangements, and investigator contact details. No incentive is offered.

The questionnaire is administered using FindMind through a non-personalised web link. No login is required. Partial responses are saved automatically, and respondents may return to incomplete questionnaires. IP addresses are not stored. Multiple submissions from the same device are permitted because hospital computers may be shared by several staff members.

Because some institutional mailing lists conceal recipient numbers, additional internal forwarding cannot be tracked, and some staff may receive the invitation more than once, the exact number of unique invitees is unknown. The minimum documented number invited will be recorded for each hospital. No conventional response rate will be calculated. An estimated participation proportion will be calculated as the number of eligible respondents divided by the minimum documented number invited and interpreted as an upper-bound estimate.

### Outcomes

#### Primary outcomes

The primary outcome group consists of one proximal self-reported behavioural indicator and three potential determinants of timely MET activation:

1. Self-reported non-activation despite fulfilment of activation criteria: agreement or strong agreement with the statement that the respondent would not call the MET when one or more MET criteria were fulfilled but the patient did not appear critically ill. This item is treated as a proximal behavioural indicator rather than as a barrier determinant.
2. Perceived pressure to avoid MET activation unless absolutely necessary (prespecified TDF domain: Social influences).
3. Concern that MET activation reflects poorly on professional competence (prespecified TDF domain: Social/professional role and identity).
4. Absence of a departmental culture supporting immediate activation when criteria are present (prespecified TDF domain: Environmental context and resources).

For binary summaries, endorsement of the first three outcomes is defined as agree or strongly agree. The positively worded culture item is coded as a barrier when respondents disagree or strongly disagree. Neutral responses are classified as non-endorsement in the main binary analysis but remain a separate category in descriptive and ordinal analyses.

#### Secondary outcomes

Secondary outcomes include individual items assessing knowledge, recall and interpretation of criteria, confidence in activation and communication, psychological safety, previous experience, perceived usefulness and effectiveness, timeliness and frequency, workload or resource barriers, fear of criticism, consultation before activation, role clarity, feedback after MET calls, assessment of local criteria, preferred activation form, and facilitators. No overall barrier, facilitator, or TDF domain score will be calculated.

The survey measures perceived timeliness rather than objectively verified delay. It does not establish that any reported determinant caused delayed activation.

### Data management and quality assurance

Data are exported from FindMind and stored on password-protected institutional computers and servers accessible only to the first and senior authors. A restricted raw dataset is preserved unchanged, and a de-identified analysis dataset is created.

The survey is confidential rather than fully anonymous. Optional contact information may be provided within the same survey record, and combinations of hospital, broad clinical area, profession, seniority, and years of employment may permit indirect identification in small subgroups. Contact information will be removed from the analysis dataset and stored separately. Participating hospitals will receive aggregated site-level results only.

Completely empty records and ineligible respondents are excluded. Partial questionnaires are retained. Duplicate records are excluded only when there is compelling evidence, such as identical contact information, an explicitly reported repeat submission, or an exact timestamp and response-pattern combination highly unlikely to occur independently. Similar demographics, rapid completion, or uniform Likert responses alone are not sufficient for exclusion. Unusually rapid submissions and straight-lined matrices are manually reviewed for additional evidence of invalid responding.

Free-text responses are reviewed for names, exact roles, locations, patient information, or other identifying content. Identifying information is removed from the analysis copy, and quotations may be lightly paraphrased when necessary to reduce deductive disclosure.

### Statistical analysis

Statistical analyses will be performed using Stata version 19 after completion of data collection at all four hospitals and finalisation of the data-cleaning log. All eligible submitted questionnaires containing at least one substantive response after the eligibility screen will be included, including partial questionnaires. Item-specific denominators will be reported, and missing outcome responses will not be imputed. Respondents with missing profession will remain in overall analyses but will be excluded from profession-specific analyses. Primary adjusted analyses will use complete cases.

Categorical variables will be reported as counts and percentages. Continuous variables will be summarised using means and standard deviations when approximately symmetric and medians and interquartile ranges otherwise. For every Likert item, the full five-category distribution will be presented. Non-substantive categories, including cannot assess, not applicable, and prefer not to answer, will be reported separately and excluded from inferential models.

The primary professional comparison is physicians versus registered nurses and nurses in training. Allied health professionals will contribute to overall descriptions and will be reported separately without primary inferential testing.

The primary adjusted analysis for each of the four prespecified primary outcomes will use ordinal logistic regression with the original five-category response. For the positively phrased departmental-culture item, response coding will be reversed so that higher values consistently indicate a less favourable determinant profile. Models will adjust a priori for hospital as a categorical fixed effect, years of professional experience, clinical area (medical, surgical, or other), and previous personal MET activation.

The proportional-odds assumption will be assessed using global and variable-specific tests and inspection of category-specific estimates. If materially violated, a partial proportional-odds model will be considered; if this is unstable or difficult to interpret, a binary logistic model using prespecified outcome endorsement will replace the ordinal model for that item. Covariates will be selected on substantive grounds, not univariable P values. Sparse categories, separation, collinearity, and model stability will be assessed. If necessary, model complexity will be reduced while retaining profession and hospital as highest-priority variables.

Secondary items will be analysed individually using full response distributions, endorsement proportions and unadjusted physician-nurse comparisons. These analyses are exploratory. No comparison between recently implemented and established MET systems will be performed because each category contains only two hospitals and MET maturity is inseparable from hospital type and other institutional characteristics.

No multiplicity correction will be applied to the four prespecified primary outcomes. Interpretation will consider effect size, confidence-interval width, consistency across outcomes, and clinical plausibility. Secondary analyses are exploratory and will not be interpreted as confirmatory based on isolated P values.

Prespecified sensitivity analyses will: (1) compare physicians with all non-physicians; (2) exclude neutral responses to compare clear endorsement with clear non-endorsement; and (3) restrict analyses to respondents reaching the final substantive questionnaire section. No post hoc power calculation will be performed.

### Qualitative analysis

Free-text responses will be analysed separately using qualitative content analysis. Categories will first be developed inductively from the data. In a second step, categories relevant to determinants of MET activation will be mapped to TDF(v2) domains where conceptually appropriate. Categories that do not fit the TDF will be retained separately. Disagreements will be resolved through discussion. Quotations will be de-identified and translated into English for publication where required.

### Sample size

The study uses a census approach by inviting all identifiable eligible staff at participating hospitals. No fixed sample-size target is imposed because the exact eligible denominator is unavailable and participation is voluntary. The feasibility and complexity of adjusted models will be determined by the number of responses and outcome events. No post hoc power calculation will be undertaken.

### Ethics and consent

The project was classified as research outside the scope of the Swiss Human Research Act. The competent ethics determination for Kantonsspital Aarau was issued by Ethikkommission Nordwestschweiz on 28 May 2025, reference Req-2025-00719.

The survey introduction explains the purpose, eligibility criteria, estimated completion time, voluntary participation, intended data use, confidentiality, and investigator contact information. Submission of responses is considered informed consent. Participants may stop at any time; however, responses entered before discontinuation may remain in the database because partial responses are saved automatically.

### Study status and protocol registration

At the time of protocol posting, data collection has been completed at one hospital and remains ongoing at three hospitals. The pooled multicentre dataset has not been analysed. This document therefore constitutes a prospective protocol and statistical analysis registration for the pooled multicentre analysis, but not a fully prospective preregistration of data collection at the first hospital.

### Dissemination

Study findings will be submitted to a peer-reviewed journal and presented at relevant clinical and scientific meetings. Participating hospitals will receive aggregated results intended to guide MET system improvement. No individual-level data will be returned.

### Patient and public involvement

Patients and members of the public were not involved in developing this staff survey because the study focuses on healthcare professionals’ implementation behaviour.

## Data Availability

The questionnaire, statistical analysis plan, and supplementary methodological materials will be publicly archived with the protocol. De-identified participant-level data will be made available only as permitted by ethics determinations, institutional policy, and disclosure-risk assessment.

## Declarations

### Ethics approval and consent to participate

Research outside the scope of the Swiss Human Research Act; site-specific determinations to be inserted. Participation is voluntary, and submission of the questionnaire indicates consent.

### Consent for publication

Not applicable.

### Authors’ contributions

NG: inception, design, data collection, manuscript draft. LC: inception, design, data collection, data analysis, manuscript draft.

## Acknowledgements

NA.

## References

1. Radeschi G, Urso F, Campagna S, et al. Factors affecting attitudes and barriers to a medical emergency team among nurses and medical doctors: a multi-centre survey. Resuscitation. 2015;88:92–98.

2. Loisa E, Hoppu S, Hytönen S-M, Tirkkonen J. Rapid response team nurses’ attitudes and barriers to the rapid response system: a multicentre survey. Acta Anaesthesiol Scand. 2021;65:695–701.

3. Szczeklik W, Fronczek J, Górka J, et al. Attitudes of healthcare professionals towards the introduction of rapid response teams in Poland: a survey study after 6 months of a pilot program in 25 hospitals. Pol Arch Intern Med. 2019;129(12):949–955.

4. Atkins L, Francis J, Islam R, et al. A guide to using the Theoretical Domains Framework of behaviour change to investigate implementation problems. Implement Sci. 2017;12:77.

